# Effect of antibiotic consumption on self-reported recovery from uncomplicated urinary tract infection

**DOI:** 10.1101/2021.03.28.21254491

**Authors:** Amal Gadalla, Hannah Wise, Kathryn Hughes, Carl Llor, Michael Moore, Theo J M Verheij, Paul Little, Chris C Butler, Nick Francis

## Abstract

**Background:** Randomised trials provide high-quality data on the effects of antibiotics for urinary tract infections (UTI) but are prone to selection bias and may not accurately reflect effects in those who actually consume antibiotics.

**Aim:** To estimate the effect of antibiotic consumption, rather than prescription, on self-reported time to recovery in women with uncomplicated UTI.

**Design and setting:** Secondary analysis of data from a randomised control trial on point of care diagnostic to guide antibiotic prescribing in primary care. Antibiotic prescribing decisions were not mandated by the trial protocol.

**Methods:** 413 adult women presenting with UTI symptoms and who either consumed (n=333) or did not consume (n=80) one or more doses of antibiotics during a 14-day follow-up were included. Antibiotic consumption was standardised using defined daily dose (DDD) and described as “days-worth of antibiotics”. Cox-proportional hazard model was used to estimate the effect of antibiotic consumption upon time to self-reported recovery, controlling for potential confounders and propensity of consuming antibiotics based on baseline symptom severity and history of UTI treatment.

**Results:** The crude median time to recovery was 3 days longer among patients who did not consume antibiotics (10 days; 95% CI= 8.5-11.5) compared to those who consumed antibiotics (7 days; 95% CI= 6.5-7.5). Patients who consumed >0 to <3, ≥3 to ≤5, and >5 days-worth of antibiotics recovered at approximately 2 – 3 times faster than the rate of those who consumed no antibiotics, adjusted HR 3.2 (95% CI 2.0, 5.1), 2.8 (1.8, 4.3), 2.3 (1.5, 3.7), respectively.

**Conclusion:** Consuming antibiotics was associated with a reduction in self-reported time to recovery in this study which is consistent with results from previous trials.

## Introduction

Urinary tract infection (UTI) is a common bacterial infection in women, affecting around half of all women over a lifetime.(1) Although uncomplicated UTI are usually easily-treated, with symptoms lasting 10 days on average,(2) they place huge financial and time burden on individuals, healthcare systems and global economy.(3-6)

Current UK guidelines for uncomplicated UTI in non-pregnant women recommend an empirical three-day antibiotic course as initial management.(7) However, strategies that reduce antibiotic prescribing and thus resistance, either with delayed prescriptions or utilizing painkillers, are being explored.(8, 9) Research shows that more than half of patients recovered with ibuprofen in the absence of antibiotics,(10) and over a third of women presenting with UTI symptoms were willing to delay antibiotic treatment.(8) However, the ever-changing situation of antibiotic resistance and perhaps the bacterial virulence of urine pathogens in community may affect the symptomatic response to treatment and patients’ willingness to avoid antibiotic treatment.(11-13)

This highlights the need for continuous investigation on the efficacy of UTI antibiotic treatment, with particular focus on including no-treatment control group. Although evidence behind current NICE guidelines for UTI treatment in non-pregnant women includes well designed randomised control trials and systematic reviews, the most recent was conducted 10 years ago. Moreover, it contains only few placebo-controlled trials and the actual documented or reported antibiotic consumption of antibiotics was rarely monitored. While placebo or a control group arm gives the opportunity to evaluate the efficacy of an antibiotic compared to the natural course of the UTI, antibiotic prescriptions will not be effective if not consumed. The aim of this study therefore is to report the effect of antibiotic consumption, as opposed to antibiotic prescription, on time to patient-reported recovery. We compared time to recovery in women who consumed variable levels of antibiotics against those who consumed no antibiotics at all.

## Methods

### Setting and participants

We utilized data collected as part of the POETIC randomised control trial (Trial number: ISRCTN65200697) that investigated the (cost-)effectiveness of a point-of-care-test guided management strategy for uncomplicated UTI between 2012 and 2014.(14) Participants were recruited from 43 general practices across England, Wales, Spain and Netherlands. General practitioners recruited adult female patients, during routine consultations, presenting with at least one of the main symptoms; dysuria, frequency or urgency. Patients were not recruited if pyelonephritis was suspected, they had received antibiotics in the four weeks prior to recruitment, were on long-term antibiotics, had significant genitourinary tract abnormalities or were terminally ill.(14)

### Treatment and follow-up of patients during POETIC study

Participating clinicians treated patients using their own judgement, or with the aid of the point of care test for those randomised to use of point-of-care-test. The test used in the study (Flexicult®) was a culture-based approach that allowed for species identification using chromogenic agar, semi-quantification, and assessment of resistance to commonly used antibiotics through the use of antibiotic-impregnated agar sections.(14) Clinicians could provide participants with an antibiotic prescription for either immediate or delayed use, or not prescribe antibiotics at all. Participants in the intervention arm had their Flexicult® result available approximately 24 hours after recruitment, and clinicians could provide participants with a change in treatment (including advising discontinuation of antibiotics, initiation of antibiotics or change in antibiotic) at this time if necessary.

A baseline questionnaire was completed by the clinician detailing patient’s symptoms and management chosen. It also reported history of previous UTI treatments. Patients were then followed up using a 14-day diary where they rated the severity of eleven symptoms (fever, pain in the side, blood in urine, smelly urine, burning or pain when passing urine [dysuria], urgency, daytime frequency, night-time frequency, tummy pain, restricted activity and general unwell feeling) on a scale of 0-6 (0= ‘not affected’ and 6= ‘as bad as can be’). Daily consumption of medication and day of recovery were also documented.

### Patients inclusion, antibiotic exposure and grouping for the current study

Data was included if a patient was prescribed no antibiotics, prescribed an antibiotic at the initial consultation with immediate effect, and had no change in antibiotic prescriptions across the 14-day follow up. Those with missing data for antibiotic use were excluded.

Patients were initially categorised into two groups based on whether they reported consuming antibiotics during follow-up or not. The group that reported consuming antibiotic were then further categorised according to the level of antibiotic consumption, based on the Defined Daily Dose (DDD) for each antibiotic.(15) This approach allows comparison between different antibiotic strengths and dosing regimens. Antibiotic consumption defined as days-worth of antibiotics was calculated by multiplying antibiotic strength (mg) by the total number of doses consumed over the treatment course (using self-reported data) and dividing it by the published DDD (the assumed averaged daily dose) for the antibiotic in question. Consumption ranged from between 0.17-14 days-worth of antibiotics and was further categorised into the following categories: >0 to <3, ≥3 to ≤5 and >5 days-worth of antibiotics.

### Statistical analysis

Kaplan-Meir survival analysis was used to estimate the median time to recovery in no antibiotic consumption groups *versus* antibiotic consumption group and *versus* different levels of antibiotic consumption. Day of recovery was censored for patients with missing recovery data or who were lost to follow up. Cox proportional hazard models were used to estimate the hazard ratio of time to recovery in antibiotic consumption groups compared to the no antibiotic consumption group.

Analgesic use (ibuprofen, paracetamol, co-codamaol, metamizole and tramadol), antifungal use (clotrimazole oral, topical and pessaries and fluconazole), anti-muscarinic use (solifenacin, trospium chloride and mebeverine hydrochloride), age and country were added to the model as covariates. A propensity score was included as a covariate in the Cox proportional hazard model to account for the probability of consuming antibiotics given the severity of the initial symptoms and UTI treatment history within the past year. The propensity score was created as the probability of antibiotic consumption in a logistic regression model with the 11 baseline symptoms scores (listed above) and UTI treatment history included as predictors. The logistic regression model with all predictors included was not significantly better than an intercept only model (−2 Loglikelihood: 281.865, χ : 15.140, P value = 0.233) and the predictors explained 7.60% of the variation in antibiotics being consumed or not (Nagelkerke Pseudo R^2^ = 0.076). Data analysis was performed using IBM SPSS Statistics.

## Results

### Participants’ characteristics and antibiotic consumption

Baseline data at the initial GP appointment were obtained from 100% of the POETIC cohort (n=643), however diary data were missing for 116 patients, who were therefore excluded. Within the first three days of starting antibiotics, 63 patients had their original antibiotic changed, and were therefore also excluded. A further 10 patients were excluded as they recorded taking a different antibiotic to the one prescribed. Patients recorded as being given a delayed antibiotic prescription (n=15) or who had missing data required to calculate levels of antibiotic consumption (n=26) were also excluded. The final cohort was 413 patients (Figure 1).

**Figure 1:**
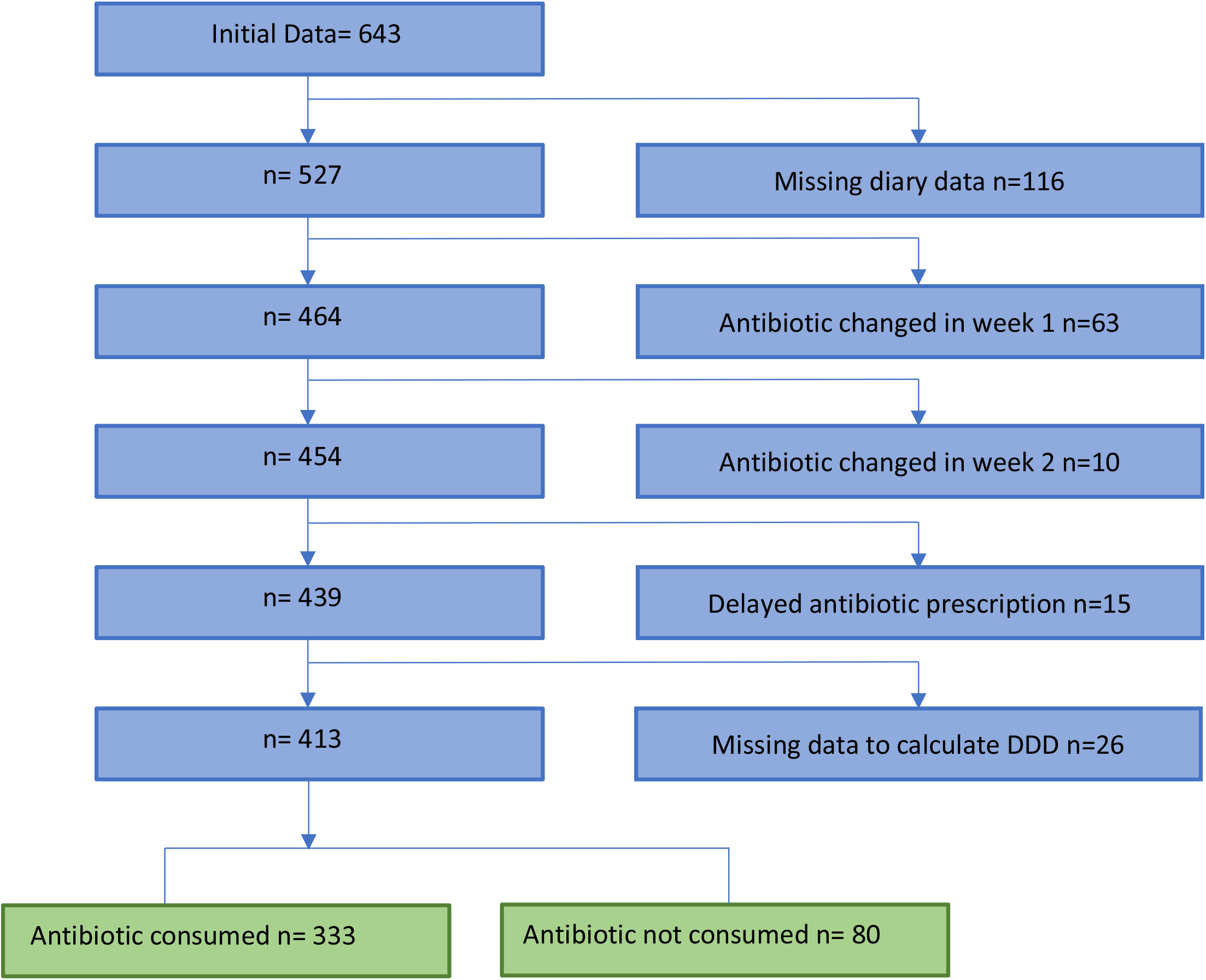
Flow chart showing application of the exclusion criteria

The no antibiotic consumption group consisted of 80 participants (75 patients were not prescribed antibiotics and did not consume any and 5 patients were prescribed antibiotics but did not consume any). Among the antibiotic-consumed group (n=333), nine different antibiotics were consumed: Trimethoprim (n=146), Nitrofurantoin (n=72), Fosfomycin (n=63), Ciprofloxacin (n=13), Norfloxacin (n=13), Amoxicillin Clavulanic (n=9), Amoxicillin (n=8), Cephalexin (n=8) and Cefuroxime (n=1).

Patients’ mean age was 49.1 (95% CI, 47.4 - 50.9) years for the entire cohort, 47.4 (95% CI, 43.4-51.4) years for the no antibiotic consumption group and 49.5 (95% CI, 47.6-51.5) years for the antibiotic consumption group (Table 1). 31.2% (129/413) of patients were from England, 33.4% (138/413) from Wales, 28.3% (117/413) from Spain and 7.0% (29/413) from Netherlands.

**Table 1:**
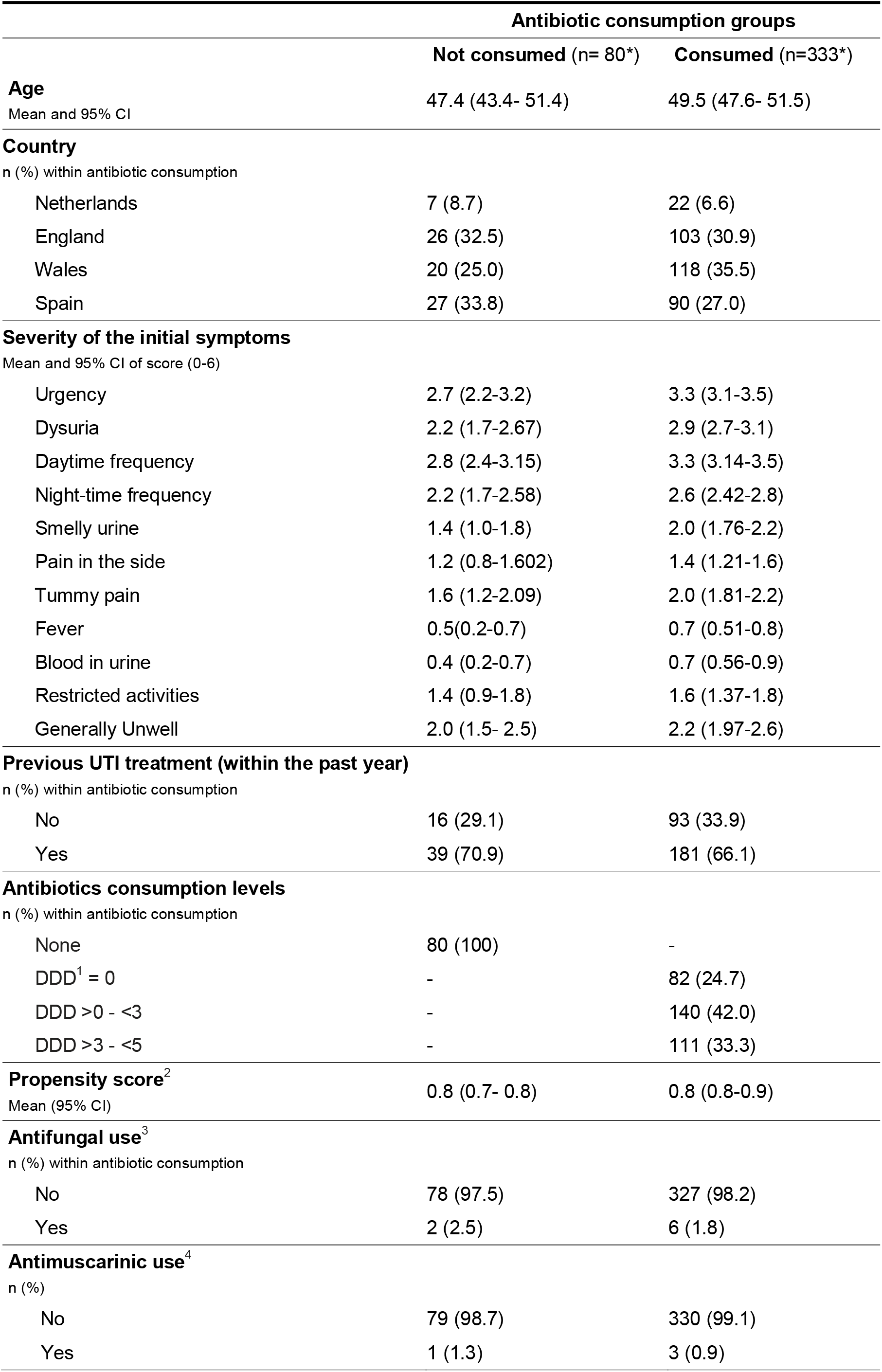
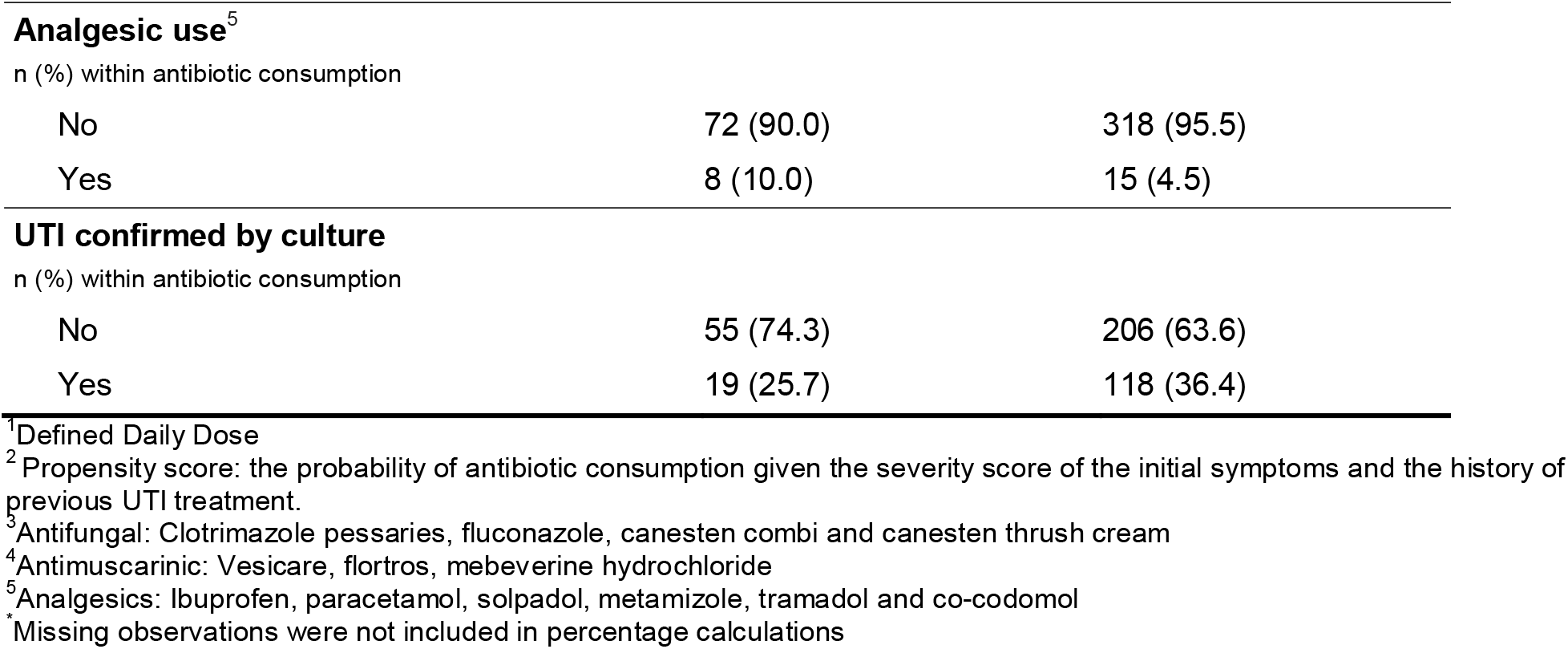
Baseline characteristics of the study group.

Severity of UTI symptoms were recorded by participants. At the baseline, 69.5% and 51.0% of the study cohort rated their daytime and night-time frequency as being ≥3, respectively. Whereas the score of ≥3 for urgency and dysuria was reported by 66.5% and 57.2% of the participants respectively. Mean symptom severity scores for daytime (P=0.006) and night-time (P=0.028) frequency, urgency (P=0.007), dysuria (P=0.006) and bad urine odour (P=0.040) were all significantly associated with antibiotic consumption. Mean scores for other symptoms were also higher in the antibiotic consumption group, but the differences were not statistically significant (Table 1). UTI treatment history within the past year was reported in 181/274 (66.1%) and 39/55 (70.9%) patients among those who did consume and did not consume antibiotics, respectively (Table 1).

Consumption of prescribed or over-the-counter analgesics, antifungals and antimuscarinics was reported at any point of the follow-up by 5.6% (23/413), 1.9% (8/413) and 1% (4/413) of participants; respectively.

There was a higher proportion of women taking analgesics (10% vs 4.5%), antifungal (2.5% vs 1.8%) or anti-muscarinic (1.3% vs 0.9%) among the no antibiotic consumed group compared to the antibiotic-consumed group, however, no statistically significant difference was detected in these percentages (Table 1). UTI was confirmed by microbiological culture following management decision was made by clinicians and there were 26% (19/74) patients with UTI positive culture among the no antibiotic consumption group and 36.4% (118/324) patients among the antibiotic consumption group (Table 1).

### Time to recovery

80 (19.4%) participants had missing data on self-reported recovery and their recovery time was therefore censored. Of those with recovery data (n=333), 331 (99.4%) reported complete recovery from UTI symptoms during the 14-day follow-up period, with only 2 (0.6%) participants reported not-recovered. Among those who consumed no antibiotics (n=80), 50 (62.5%) participants reported recovery during the follow-up period, the remaining participants had censored recovery time. Among participants who consumed antibiotics (n=333), 281 (84.4%) participants reported recovery during the follow-up period and the remaining participants had censored recovery time.

The overall estimated median time at which 50% of the cohort had recovered was 8.0 (95% CI 7.5 – 8.5) days. The crude estimated median time to recovery was 3 days longer among participants who did not consume any antibiotics (10 days; 95% CI= 8.5-11.5, Log rank test P < 0.001) compared to those who consumed antibiotics (7 days; 95% CI= 6.5-7.5), Figure 2A.

**Figure 2:**
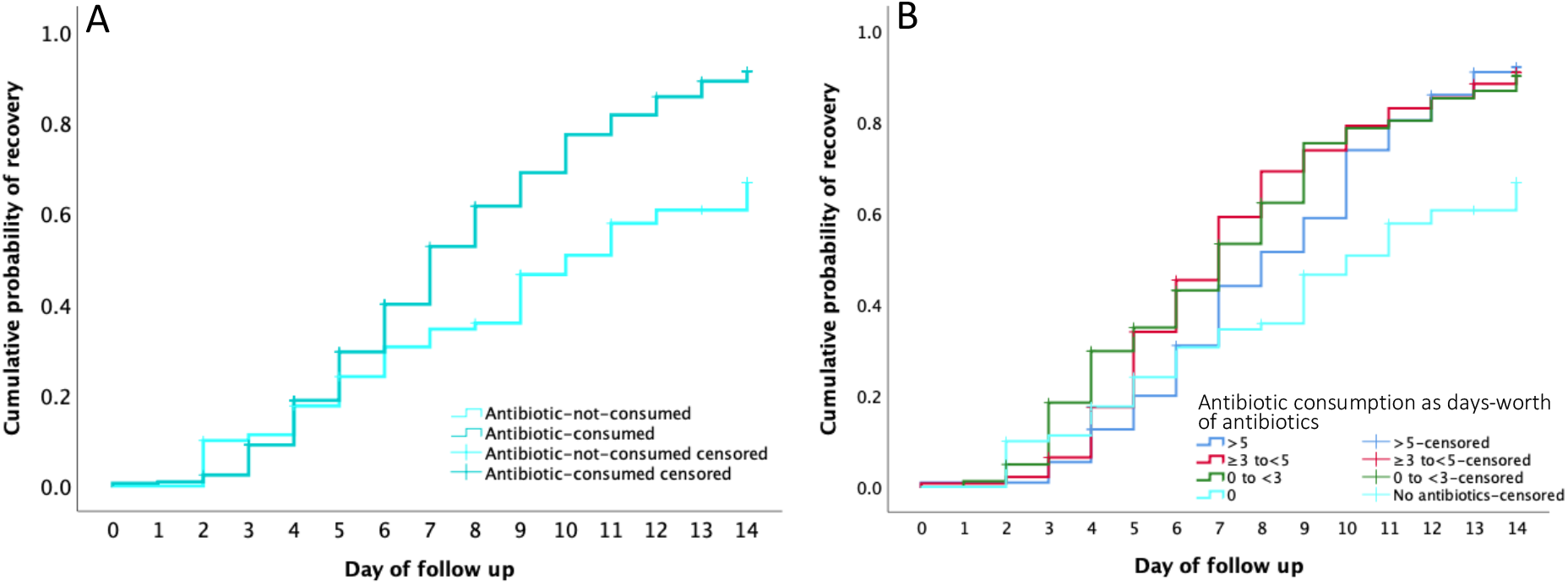
Crude median time to recovery. **(A)** across antibiotic consumption and no-antibiotic consumption. **(B)** across no antibiotic consumptions and variable levels of antibiotic consumption.

There were 82 (24.6%), 140 (42.1%) and 111 (33.3%) participants who consumed >0 to <3, ≥3 to ≤5 and >5 days-worth of antibiotics, respectively. The crude estimated median time to recovery was significantly different (Log Rank test, χ^2^= 23.2, P < 0.001) for those who consumed no antibiotics (10 days; 95% CI= 8.5-11.5), consumed >0 to <3 days-worth of antibiotics (7 days; 95% CI = 5.8 - 8.2), ≥3 to ≤5 days-worth of antibiotics (7 days; 95% CI = 6.3 - 7.7), or >5 days-worth of antibiotics were (8 days; 95% CI = 6.7 - 9.3), Figure 2B. In a univariate Cox-proportional model, each of the antibiotic consumption levels recovered at approximately 2 – 3 times faster than the rate of those who did not consume antibiotics (Table 2). After adjusting for analgesic use, antifungal use, antimuscarinic use, age, country and propensity score based on initial symptoms and history of previous UTI, the hazard ratios increased (Table 2).

**Table 2.** Hazard ratio of recovery among antibiotic consumption levels compared to no antibiotic consumption.

|  | Hazard Ratio | 95% CI<sup>1</sup> |  |
| --- | --- | --- | --- |
|  |  | Lower | Upper |
| Univariate model |  |  |  |
| Antibiotics consumption |  |  |  |
| None |  | Referent |  |
| >0 - ≤3 days | 2.03 | 1.40 | 2.95 |
| >3 - ≤5 days | 2.03 | 1.45 | 2.83 |
| >5 days | 1.71 | 1.22 | 2.41 |
| Multivariate model<sup>*</sup> |  |  |  |
| Antibiotics consumption |  |  |  |
| None |  | Referent |  |
| >0 - ≤3 days | 3.21 | 2.0 | 5.14 |
| >3 - ≤5 days | 2.76 | 1.78 | 4.28 |
| >5 days | 2.34 | 1.50 | 3.66 |
| Propensity score<sup>3</sup> | 0.17 | 0.04 | 0.77 |
| Age | 0.99 | 0.98 | 1.00 |
| Country<sup>4</sup>: |  |  |  |
| Netherlands |  | Referent |  |
| England | 1.04 | 0.58 | 1.89 |
| Wales | 0.86 | 0.49 | 1.50 |
| Spain | 0.78 | 0.43 | 1.42 |
| Antifungal use<sup>5</sup> |  |  |  |
| Yes |  | Referent |  |
| No | 1.53 | 0.62 | 3.79 |
| Antimuscarinic use<sup>6</sup> |  |  |  |
| Yes |  | Referent |  |
| No | 0.96 | 0.23 | 3.93 |
| Analgesic use<sup>7</sup> |  |  |  |
| Yes |  | Referent |  |
| No | 1.79 | 0.99 | 3.22 |

<sup>1</sup> 95% Confidence intervals
<sup>2</sup> Defined Daily Dose
<sup>3</sup> Propensity score: the probability of antibiotic consumption given the severity score of the initial symptoms and the history of previous UTI treatment.
<sup>4</sup> Countries: participants were from England, Wales, Spain and Netherland
<sup>5</sup> Anti-fungal: Clotrimazole pessaries, fluconazole, canesten combi and canesten thrush cream
<sup>6</sup> Anti-muscarinic: Vesicare, flortros, mebeverine hydrochloride
<sup>7</sup> Analgesics: Ibuprofen, paracetamol, solpadol, metamizole, tramadol and co-codamol
<sup>\*</sup>Significant reduction in -2 Log Likelihood between the univariate (2596.9) and multivariate (2578.9) Cox-proportional models ( $\chi^2=17.97$ , df=8, P = 0.021475)

## Discussion

### Summary

In this secondary data analysis from an international trial of a point-of-care-test to guide the management of UTI, we found that antibiotic consumption reduced time to recovery from UTI symptoms. Participants who consumed >0 to <3 days of antibiotics, ≥3 to ≤5 days of antibiotics, and >5 days of antibiotics all recovered at about 2 – 3 times faster than the rate of those who did not consume antibiotics, after controlling for baseline characteristics, use of other medication, and propensity to be prescribed antibiotics. There were no important differences between the different levels of antibiotic consumption.

### Strengths and limitations

To our knowledge, only one other study (10) explicitly explored recovery from UTI symptoms using antibiotic consumption data rather than antibiotic prescriptions. Non-adherence is a recognised issue in general practice for all infections.(16) There were five patients in our study who were prescribed antibiotics but did not report consuming any. Our study included all of the antibiotics most commonly used in primary care to treat uncomplicated UTI, and we standardised variable antibiotic exposure (type, dose and strength) by calculating the number of days-worth of antibiotic consumption based on the DDD.

A further strength of this study is use of propensity scores of initial symptoms and the history of previous UTI treatment as a covariate in estimating hazard ratio of recovery. Such an approach helps correct for indication bias, where those with more severe symptoms are more likely to be prescribed and consume antibiotics. We have also corrected for the potential confounders: age, country, and analgesic, antifungal and antimuscarinic use.

The main limitation of this study is risk of residual confounding from unmeasured confounders such as signs that could make GPs not to prescribe antibiotics. In addition, data for comorbidity were not considered as it was not recorded in the POETIC. There were no data collected on the duration of the current UTI symptoms, but the study did not include patients with UTI symptoms for longer than 14 days. Although antimuscarinic, antifungal and analgesic use was adjusted for, we used binary indicators and did not take into account the amount, duration or time point these were consumed, as this information was not collected. Nevertheless, none of these variables had a significant effect, or changed the hazard ratio for recovery, when adjusted for in the models. We excluded patients who changed antibiotics during the follow-up period or who reported consuming a different antibiotic from the one that they had been prescribed. This was to limit potential bias, for example antibiotics might be changed because of severe symptoms or evidence of resistance, which could affect outcomes. A relatively small number of patients will fall into this category but by excluding them we may have introduced unintended bias. Finally, we used antibiotic consumption provided by patients, which may not be as valid as more objective measures such as medication adherence monitoring containers or measurement of antibiotic levels in blood or urine. (17)

### Comparison with existing literature

The most recent Cochrane review comparing different antibiotics for UTI management found that most antibiotics effectively treated uncomplicated UTI symptoms, however lack of data prevented evaluation of time to symptomatic recovery.(18) The evidence from placebo-controlled trials is scarce. Hoffmann et al. reviewed the natural course of uncomplicated UTI in women on studies up to November 2019, they found only three placebo-controlled randomized trials and reported symptoms duration.(19) Their results showed that complete symptom recovery could occur in 18% (up to 3 days) to 54% (up to 6 weeks) of the placebo arm, with most improvement occurring in the first 9 days. These findings are in line with our results as we found symptoms were completely resolved within the study follow up in nearly two thirds (62%) of the women who did not consume antibiotics.

Moreover, antibiotics were found to be superior to no antibiotic management strategies, such as painkillers, for uncomplicated UTI in women. Vik et al. found that while ∼40% of women recovered by day 4 with ibuprofen, ∼70% recovered with pivmecillinam.(20) With higher dose of ibuprofen, Gágyor et al. found that 70% of women were recovered by day 7 compared to 82% for fosfomycin. They concluded no antibiotic management would apply mostly to women with mild to moderate symptoms.(10) Another study that compared diclofenac *versus* norfloxacin found that median time to recovery with diclofenac was 4 days compared to 2 days for norfloxacin.(21) These findings are in line with our study as women consumed no antibiotics took longer to recover.

### Implications for research and/or practice

Our study found an association between consumption of antibiotics and shorter time to recovery in women with uncomplicated UTI in primary care. This finding is consistent with evidence from trials suggesting net overall benefit from antibiotic use in women with uncomplicated UTI. Due to the nature of the design of this observational study, we are not able to conclude that the use of antibiotics *causes* a faster rate of recovery. Although we attempted to control for baseline symptom severity and other potential confounders, it is possible that the association observed in this study is because women who were less likely to consume antibiotics were more likely to have a prolonged recovery (if they had atypical symptoms for example). However, it is clear that many women with symptoms of UTI do make a good recovery without antibiotic treatment, and attention should focus on trying to identify these women. It is possible that some of them do not actually have bacterial infections (culture confirmed bacterial UTI in 25.7% and 36.4% among the no consumption and consumption groups, respectively), or that their host defences are able to cause resolution without the need for antibiotics. There is also a need for further studies quantifying the risk and predictors of complications and adverse effects associated with UTI managed with and without antibiotic treatment.

If patients were aware of the likelihood of reaching recovery by day 14 without antibiotics, some may choose to refrain from immediate antibiotics, which supports delayed antibiotic strategies. This may be particularly useful in younger patients and in those with lower risk for pyelonephritis (8, 21, 22) and could lead to a reduction in antibiotic use and risk of antibiotic resistance as evident in respiratory tract infections.(23)

## Data Availability

Anonymised data will be available upon request. The corresponding author or the senior authors (Nick Francis: and Chris Butler) can receive email requests.

## Funding

This secondary analysis was supported by Cardiff University Intercalated BSc in Population Medicine Program 2018/2019

## Ethical approval

POETIC was approved in the UK by the Research Ethics Committee for Wales (now known as Wales REC3) (reference: 12/WA/0394), Jordi Gol i Gurina Ethics Committee in Barcelona (reference: AC13/01), and the Medical Research Ethics Committee of UMC Utrecht (reference: 13/304) in the Netherlands. The use of data for future studies concerning UTI diagnosis and management was consented for during the POETIC study. All patients’ data were completely anonymised for the current study research team.

## Acknowledgments

This research was partially supported by the Division of Population of Medicine, Cardiff University.

## Notes

### Competing Interest Statement

The authors have declared no competing interest, except Dr. Francis reports grants from European Union FP7, during the conduct of the study; personal fees and non-financial support from Abbott, outside the submitted work. Dr. Llor reports grants from Abbott Diagnostics, outside the submitted work.

### Author Declarations

POETIC was approved in the UK by the Research Ethics Committee for Wales (now known as Wales REC3) (reference: 12/WA/0394). Jordi Gol i Gurina Ethics Committee in Barcelona (reference: AC13/01) and the Medical Research Ethics Committee of UMC Utrecht (reference: 13/304) in the Netherlands. The use of data for future studies concerning UTI diagnosis and management was consented for during the POETIC study. All patients data were completely anonymised for the current study research team.

## References

1. Foxman B. Epidemiology of urinary tract infections: incidence, morbidity, and economic costs. Disease-a-month : DM. 2003;49(2):53–70.

2. Ferry SA, Holm SE, Stenlund H, Lundholm R, Monsen TJ. The natural course of uncomplicated lower urinary tract infection in women illustrated by a randomized placebo controlled study. Scand J Infect Dis. 2004;36(4):296–301.

3. Dolk FCK, Pouwels KB, Smith DRM, Robotham JV, Smieszek T. Antibiotics in primary care in England: which antibiotics are prescribed and for which conditions? Journal of Antimicrobial Chemotherapy. 2018;73(suppl_2):ii2–ii10.

4. Foxman B, Barlow R, D’Arcy H, Gillespie B, Sobel JD. Urinary tract infection: self-reported incidence and associated costs. Ann Epidemiol. 2000;10(8):509–15.

5. Foxman B, Frerichs RR. Epidemiology of urinary tract infection: II. Diet, clothing, and urination habits. Am J Public Health. 1985;75(11):1314–7.

6. Gonzalez CM, Schaeffer AJ. Treatment of urinary tract infection: what’s old, what’s new, and what works. World J Urol. 1999;17(6):372–82.

7. NICE. Urinary Tract Infection (lower)- women 2015 [Available from: https://cks.nice.org.uk/urinary-tract-infection-lower-women#!scenario.

8. Knottnerus BJ, Geerlings SE, Moll van Charante EP, ter Riet G. Women with symptoms of uncomplicated urinary tract infection are often willing to delay antibiotic treatment: a prospective cohort study. BMC Fam Pract. 2013;14:71.

9. Little P, Moore M, Kelly J, Williamson I, Leydon G, McDermott L, et al. Delayed antibiotic prescribing strategies for respiratory tract infections in primary care: pragmatic, factorial, randomised controlled trial. BMJ : British Medical Journal. 2014;348:g1606.

10. Gágyor I, Bleidorn J, Kochen MM, Schmiemann G, Wegscheider K, Hummers-Pradier E. Ibuprofen versus fosfomycin for uncomplicated urinary tract infection in women: randomised controlled trial. BMJ. 2015;351:h6544.

11. Abbo LM, Hooton TM. Antimicrobial Stewardship and Urinary Tract Infections. Antibiotics (Basel). 2014;3(2):174–92.

12. Bush K, Courvalin P, Dantas G, Davies J, Eisenstein B, Huovinen P, et al. Tackling antibiotic resistance. Nature Reviews Microbiology. 2011;9(12):894–6.

13. Pujades-Rodriguez M, West RM, Wilcox MH, Sandoe J. Lower Urinary Tract Infections: Management, Outcomes and Risk Factors for Antibiotic Re-prescription in Primary Care. EClinicalMedicine. 2019;14:23–31.

14. Bates J, Thomas-Jones E, Pickles T, Kirby N, Gal M, Bongard E, et al. Point of care testing for urinary tract infection in primary care (POETIC): protocol for a randomised controlled trial of the clinical and cost effectiveness of FLEXICULT(tm) informed management of uncomplicated UTI in primary care. BMC Family Practice. 2014;15(1):187.

15. WHO. Defined Daily Dose (DDD): World Health Organizaion; 1990 [cited 2021. Available from: https://www.who.int/tools/atc-ddd-toolkit/about-ddd.

16. Francis NA, Gillespie D, Nuttall J, Hood K, Little P, Verheij T, et al. Antibiotics for acute cough: an international observational study of patient adherence in primary care. The British journal of general practice : the journal of the Royal College of General Practitioners. 2012;62(599):e429–37.

17. Czobor P, Skolnick P. The secrets of a successful clinical trial: compliance, compliance, and compliance. Mol Interv. 2011;11(2):107–10.

18. Zalmanovici Trestioreanu A, Green H, Paul M, Yaphe J, Leibovici L. Antimicrobial agents for treating uncomplicated urinary tract infection in women. Cochrane Database Syst Rev. 2010(10):Cd007182.

19. Hoffmann T, Peiris R, Mar CD, Cleo G, Glasziou P. Natural history of uncomplicated urinary tract infection without antibiotics: a systematic review. British Journal of General Practice. 2020;70(699):e714–e22.

20. Vik I, Bollestad M, Grude N, Bærheim A, Damsgaard E, Neumark T, et al. Ibuprofen versus pivmecillinam for uncomplicated urinary tract infection in women-A double-blind, randomized non-inferiority trial. PLoS Med. 2018;15(5):e1002569.

21. Kronenberg A, Bütikofer L, Odutayo A, Mühlemann K, da Costa BR, Battaglia M, et al. Symptomatic treatment of uncomplicated lower urinary tract infections in the ambulatory setting: randomised, double blind trial. BMJ. 2017;359:j4784.

22. Bjerrum L, Lindbæk M. Which treatment strategy for women with symptoms of urinary tract infection? BMJ. 2015;351:h6888.

23. Francis NA, Gillespie D, Nuttall J, Hood K, Little P, Verheij T, et al. Antibiotics for acute cough: an international observational study of patient adherence in primary care. British Journal of General Practice. 2012;62(599):e429–e37.

